# Empty Streets, Speeding and Motor Vehicle Collisions during Covid-19 Lockdowns: Evidence from Northern Ireland

**DOI:** 10.1101/2021.01.03.21249173

**Authors:** Sotiris Vandoros, Fotis Papailias

## Abstract

Covid-19 and lockdowns have had spillover effects on other health outcomes. Motor vehicle collisions (MVC) are likely to have been affected by the pandemic due to, among others, less traffic volume and speeding on empty streets. This paper studies the impact of the pandemic on MVCs in Northern Ireland. Using monthly data on injuries and deaths, we find a steep decline in slight and serious injuries compared to what would have been expected in the absence of the pandemic. However, we find no effect on the number of deaths. Based on data from speeding tickets, a plausible explanation for the differential effect on the number of injuries and deaths is speeding on empty streets during the pandemic.

## 1. Background

Over 50 people lose their lives in Motor Vehicle Collisions (MVC) in Northern Ireland every year, and more than 700 are seriously injured.^1^ As Covid-19 has been affecting nearly all aspects of everyday activities, there are reasons to believe that the number of MVCs has also been affected by the pandemic, as outlined by plausible mechanisms put forward in a previous study.^2^

First, the impact on traffic volume may have an ambiguous effect on the number and severity of MVCs. People are likely to commute less due to lockdowns and due to being afraid of catching Covid-19. On one hand, less traffic volume means fewer vehicles at risk of collision. Higher unemployment rates observed during the pandemic^3^ also are likely to lead to a reduction in the number of collisions, as the experience from previous recessions has shown.^4-5^On the other hand, less congested streets give an opportunity for speeding, which may affect the likelihood and severity of collision.^6-7^

Second, factors associated with the pandemic have had an impact on driving behaviour. Distraction, due to worrying about family members who are ill^8^ or due to economic uncertainty and financial worries,^9-11^ has been associated with increased risk of MVC. Changes in drinking patterns have also occurred during the pandemic. Bars and restaurants were closed for certain periods, due to measures to tackle Covid-19, and had fewer customers when open as people were trying to reduce the risk of catching the virus. However, overall alcohol consumption may have increased.^12^ Sleeping patterns (another risk factor) have also been affected by the pandemic.^13-14^

Empirical evidence on the impact of the Covid-19 pandemic on MVCs is scarce, as many countries publish data with a significant time lag. Early empirical evidence suggests that there has been a decrease in collisions during the pandemic.^2,15-18^ Interestingly, in the USA there has been a reduction in MVCs in the first months of the Covid-19 pandemic,^15-16^ but at the same time there has been an increase in deaths.^19^

The objective of this study is to examine whether and to what extent MVCs, injuries and deaths were affected by the Covid-19 pandemic in Northern Ireland.

## 2. Data and Methods

We obtained monthly data on MVCs from the Police Service of Northern Ireland (PSNI). Data included the monthly number of slight injuries, serious injuries and deaths for the period from January 2015 to October 2020. Unfortunately, monthly data by cause of MVC were not available, but the PSNI provides monthly data on the number of traffic-related fines by type of violation.

We investigated the lockdown effects employing Interrupted Time Series (ITS) analysis. The “intervention” started in March, when the first Covid-19 death occurred in Northern Ireland, and the first lockdown was introduced. This is also the period when reduced mobility was observed.^20^ Separate analyses were conducted depending on the MVC outcome: number of slight injuries, serious injuries, and deaths. The model includes a time variable, a post-intervention dummy variable, and an interaction between the post-intervention dummy and the time variable, to capture any change in trend. In particular, for each target series we estimated the model presented in Equation (1).

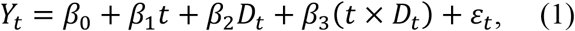

where *t* is the time trend, *D*_*t*_ is a dummy variable which takes the value 0 in the pre-lockdown period and the value 1 in the post-lockdown period and *ε*_*t*_ is the error term assuming the standard assumptions hold. *β*_0_ can be interpreted as the overall (baseline) level at time *t* = 0, *β*_1_ can be interpreted as the pre-lockdown trend, *β*_2_ indicates the change in the level after the lockdown and *β*_3_ indicates the change in the slope. The above model can be estimated assuming Gaussianity or Poisson (which is the standard approach in population health evaluations where the outcome is a count variable). The extended set of our results includes both approaches as well as the Quasi Poisson which accounts for overdispersion, even if this does not seem to be the case with our data.

## 3. Results

### 3.1 Results on the number of collisions, injuries and deaths

Figure 1 shows trends in MVCs in 2020 and previous years. There is a clear steep decline in the number of slight injuries in March and April 2020 compared to the same months in previous years (panel A). The number of slight injuries started increasing after the lockdown but remained below that of previous years throughout the study period. There is also a decrease in the number of serious injuries, although the difference compared to previous years is not as strong as in the case of slight injuries (panel B). Although in January 2020 the number of serious injuries was larger than that in any of the previous five years, this figure remained much lower than previous years’ levels during the first lockdown, and below or at the lower end of the distribution in subsequent months. However, there is little or no evidence of any decrease in deaths during the first lockdown or in the following months (panel C).

**Figure 1.**
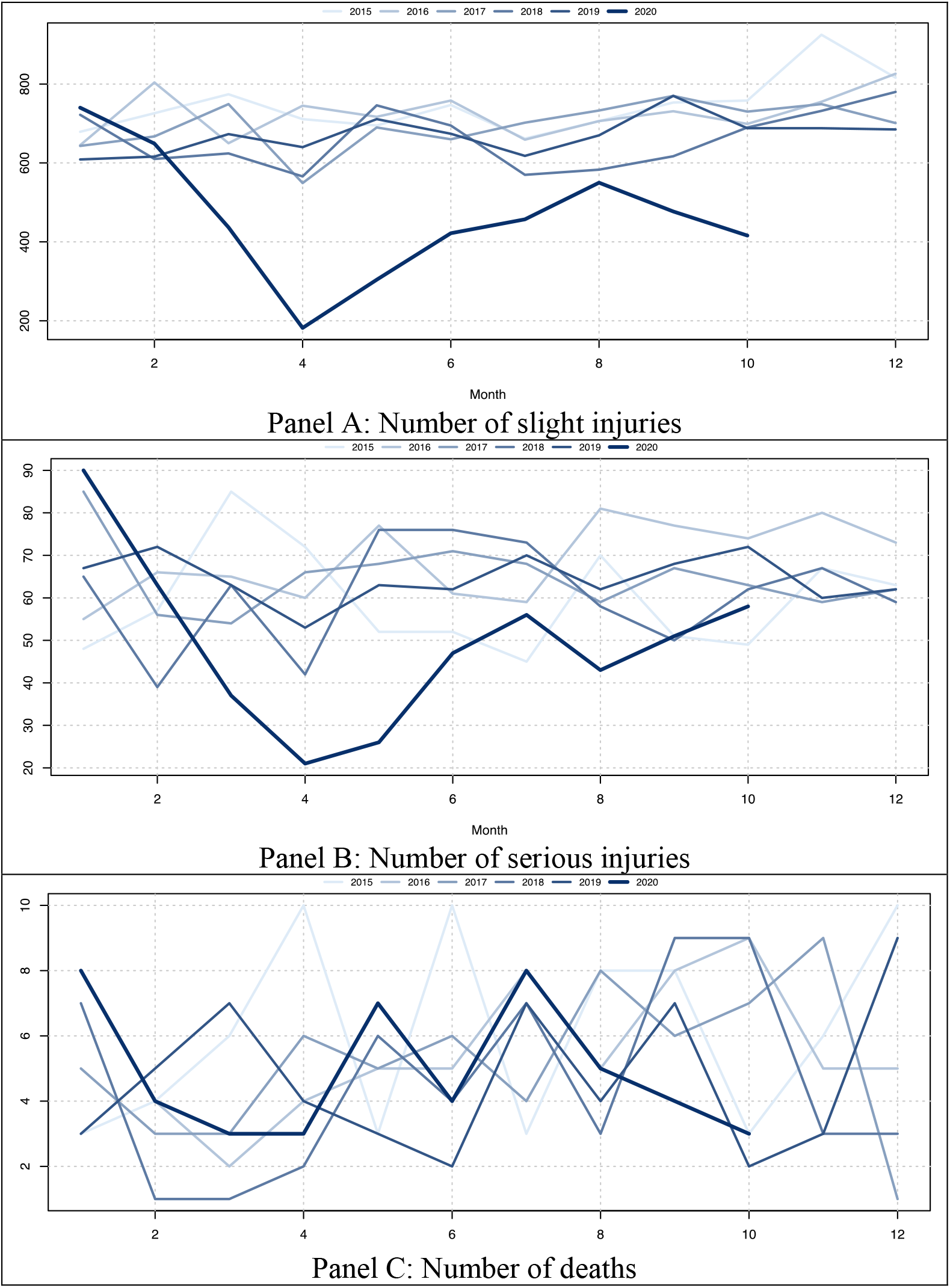
Trends in Motor Vehicle Collisions in 2020 and previous five years

We naturally divided our sample in the pre-lockdown period (from January 2015 to February 2020) and the post-lockdown period (from March 2020 to October 2020) and started our exploratory data analysis (EDA) by investigating a simple time scatterplot for the three underlying target series. A first look reveals that there seems to be a slight, and possibly not statistically significant, break in the trend and a change in the direction of the slope for deaths (*Y*_1_). This is more evident for serious injuries (*Y*_2_) and slight injuries (*Y*_3_) where there is a downward shift in the entire line, with a simultaneous change in the slope (Figure 2).

**Figure 2.**
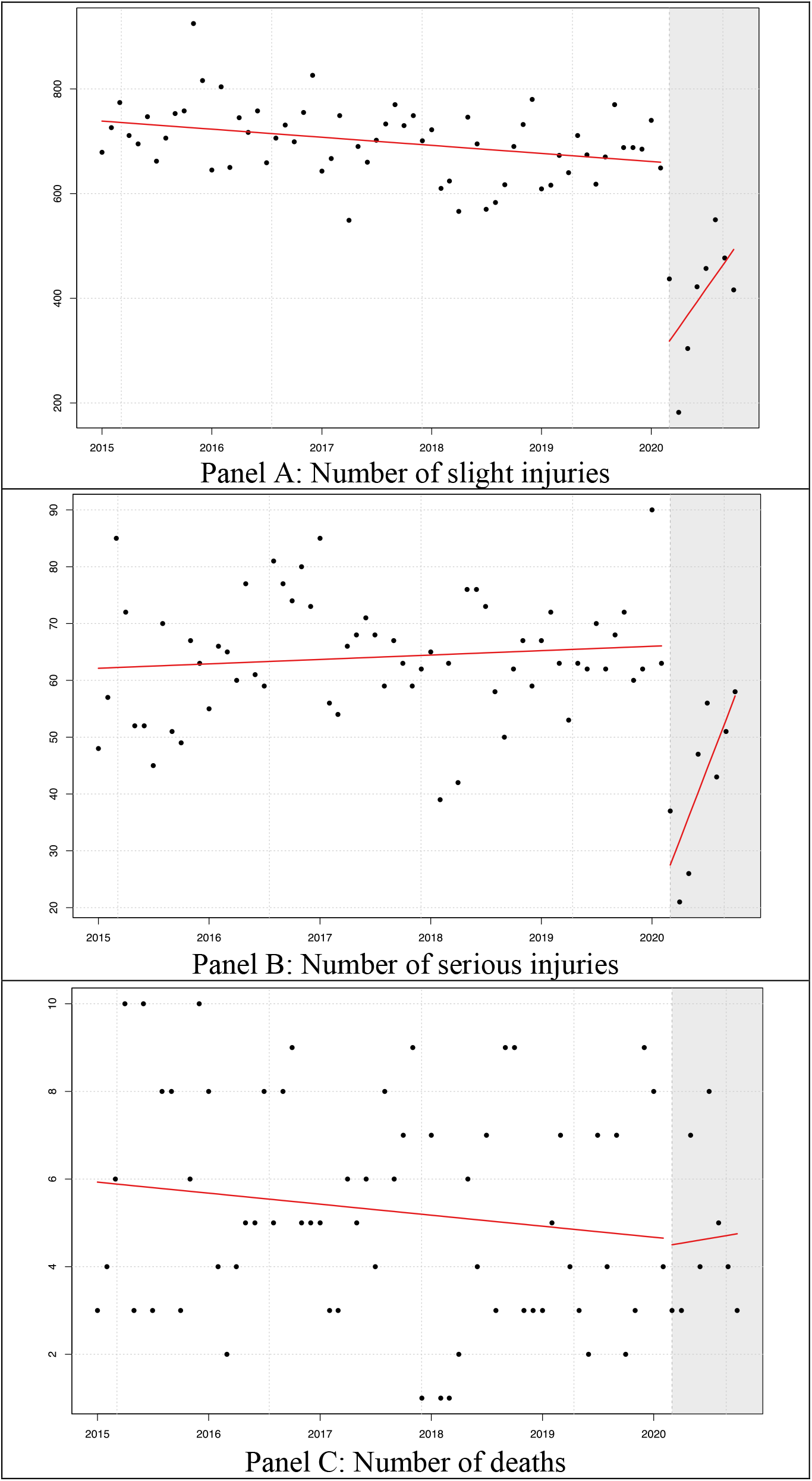
Scatterplot and trends: Motor Vehicle Collisions in 2020 and previous five years

We extended our EDA by calculating the descriptive statistics for the full sample, as well as the pre- and post- lockdown subsamples (Table 1). Table 1 includes various statistics evaluated across the full, pre- and post- lockdown samples. We compared the means of the pre- and post-lockdown samples employing a T-Test allowing for unequal variances. Even this simplified approach indicates that there is a statistically significant decrease in the average level of serious and slight injuries. In particular, serious injuries decreased by 33.89% on average (from 64.097 to 42.375; *p*-value=0.002) and slight injuries decreased by 41.99% on average (from 699.29 to 405.625; *p*-value=0.000). However, there is no statistically significant change in the number of deaths (*p*-value=0.396).

**Table 1.** Descriptive Statistics

| | Deaths ($Y_1$) | | | Serious Injuries ($Y_2$) | | | Slight Injuries ($Y_3$) | | |
| --- | --- | --- | --- | --- | --- | --- | --- | --- | --- |
|  | Full | Pre | Post | Full | Pre | Post | Full | Pre | Post |
| **Observations** | 70 | 62 | 8 | 70 | 62 | 8 | 70 | 62 | 8 |
| **Sample Mean** | 5.214 | 5.290 | 4.625 | 61.614 | 64.097 | 42.375 | 665.729 | 699.290 | 405.625 |
| **T-Test (pval)** |  | 0.396 |  |  | **0.002** |  |  | **0.000** |  |
| **Std. Dev.** | 2.443 | 2.505 | 1.923 | 12.765 | 10.417 | 13.522 | 119.376 | 67.986 | 113.562 |
| **Min** | 1 | 1 | 3 | 21 | 39 | 21 | 182 | 549 | 182 |
| **Median** | 5 | 5 | 4 | 62.5 | 63 | 45 | 690 | 700 | 429.5 |
| **Max** | 10 | 10 | 8 | 90 | 90 | 58 | 925 | 925 | 550 |
| **Skewness** | 0.272 | 0.204 | 0.678 | -0.588 | 0.033 | -0.365 | -1.544 | 0.273 | -0.742 |
| **Kurtosis** | -1.030 | -1.079 | -1.328 | 0.929 | -0.047 | -1.573 | 3.547 | 0.775 | -0.688 |

Table 2 presents the ITS results for the three dependent variables. There is a simultaneous change in the trend as well as the slope in the post-lockdown periods for serious injuries (Y_2_) and slight injuries (Y_3_). In accordance with the results of the T-Test, there is no statistically significant difference for the Deaths (*Y*_1_) series. However, we have a negative change in the level after the lockdown (*β*_2_) and an abrupt increase in the slope (*β*_3_) for the Serious Injuries (*Y*_2_) and Slight Injuries (*Y*_3_) series.

**Table 2.** Results of the interrupted time series analysis – Gaussian

| | $Y_1$ | $Y_2$ | $Y_3$ |
| --- | --- | --- | --- |
| $\hat{\beta}_0$ | 5.950<br>[4.687, 7.214]<br>**p=0.000** | 62.063<br>[56.755, 67.371]<br>**p=0.000** | 739.866<br>[704.515, 775.216]<br>**p=0.000** |
| $\hat{\beta}_1$ | -0.021<br>[-0.056, 0.014]<br>p=0.235 | 0.065<br>[-0.082, 0.211]<br>p=0.382 | -1.288<br>[-2.264, -0.312]<br>**p=0.010** |
| $\hat{\beta}_2$ | -3.700<br>[-54.174, 46.774]<br>p=0.884 | -302.313<br>[-514.364, -90.262]<br>**p=0.006** | -1997.532<br>[-3409.659, -585.406]<br>**p=0.006** |
| $\hat{\beta}_3$ | 0.057<br>[-0.702, 0.816]<br>p=0.882 | 4.185<br>[0.996, 7.375]<br>**p=0.011** | 26.300<br>[5.062, 47.538]<br>**p=0.016** |
| N | 70 | 70 | 70 |
| R<sup>2</sup> | 0.029 | 0.372 | 0.682 |

Figure A1 in the Appendix illustrates the autocorrelograms of the residuals from the regression of each series. It is evident that there is no autocorrelation or obvious seasonality patterns.

As an additional check, we also calculated an extended set of results including: Newey-West HAC t-values and confidence intervals;^21^ Poisson distribution for the error term; and Quasi Poisson Distribution to account for overdispersion. Results are presented in Tables A1, A2 and A3 respectively, in the Appendix. In all cases, our qualitative conclusions about *β*_2_and *β*_3_ still hold and confirm the results of the baseline ITS model.

### 3.2 The potential role of speeding on empty streets

Interestingly, while there was a steep decline in the number of injuries, there didn’t seem to be any change in the number of deaths. Could this be a result of greater collision severity, due to speeding on empty streets?^6-7^ Or could it be a result of increased alcohol consumption?^12^ Figure A2 in the Appendix shows the trends in speeding fines and fines relating to driving under the influence of alcohol or drugs in 2020 and previous years (before 2017 there did not seem to be much testing for alcohol). There is a steep increase in speeding fines in May and June 2020 compared to years 2017-2019. This could be a result of increased traffic violations, or a result of a greater police crackdown as a result of more speeding-related accidents. There didn’t seem to be any relative increase in the number of alcohol and drug violations, possibly because people were drinking more at home, as bars and restaurants were shut, or because they were afraid of catching the virus there after they opened.

## 4. Discussion

This study shows that there has been an increase in MVC-related injuries in Northern Ireland during the Covid-19 pandemic. There was a decrease in slight injuries in March-October 2020 (which largely coincided with the first lockdown and other social distancing measures) by about 42%, reflecting reduced mobility during and after the lockdown.^20^ Serious injuries demonstrated a reduction of about 34% over the same period. However, there was no statistically significant change in the number of deaths as a result of collision. Interestingly, while the total number of collisions was reduced, roughly the same number of people died as would have been expected in the absence of the pandemic – raising interesting questions about collision severity and reduced collision numbers.

This differential impact on the number of injuries and the number of deaths is likely to be a result of speeding in less congested streets, due to reduced traffic volume during the lockdown and the pandemic in general. Unfortunately, monthly data by cause of MVC were not available. However, an increase in fines for speeding in May and June 2020 points in that direction. Of course, the number of fines is affected by policing intensity – but even if there was a crackdown on speeding particularly in May-June 2020, this would likely be a response to increased speeding violations. Speeding can lead to more severe crashes, thus resulting in death. Therefore, it appears that the negative impact of the reduced number miles travelled was counterbalanced by higher speeds.

Another factor that may have not allowed the number of deaths to decrease is health system capacity during the pandemic. As ambulances and hospitals were responding to large numbers of Covid-19 patients, this may have led to a delay in treatment of MVC injuries, that could lead to more fatalities that otherwise. However, it is important to note that this is just a plausible explanation and this does not follow directly from the findings of this study. Future research can examine whether health system capacity was in any way related to MVC fatalities during the pandemic.

An increase in alcohol consumption reported elsewhere during the pandemic could also have been a factor,^12^ but there was no evidence of an increase in fines relating to drunk driving. It is possible that increased alcohol consumption was counterbalanced by drinking at home rather than in pubs or restaurants, thus avoiding drink driving. It is not possible to examine the role of any change in sleep duration and patterns with the data currently available. This can be examined by future research.

Results of this study add to existing research on how the pandemic affected MVCs. While there is consensus on a reduction in collisions,^15-18^ the results on fatalities are not always in the same direction.^2,19^

Findings also provide additional evidence on the spillover effects of Covid-19 on other health outcomes in Northern Ireland and elsewhere.^22-27^ Overall, this study highlights yet another indirect consequence of the pandemic in the short run, but as people adapt their behaviour, additional evidence will be needed to study the long-term effects on motor vehicle collisions.

## Data Availability

The data used on collisions are freely available on the website of the Police Service of Northern Ireland.

## Ethics approval

The data used were aggregate anonymous data, so ethics approval was not required.

## Checklist

There is no relevant checklist of observational studies.

## Appendix

**Table A1.** Gaussian regression with HAC confidence intervals

| | $Y_1$ | $Y_2$ | $Y_3$ |
| --- | --- | --- | --- |
| $\hat{\beta}_0$ | 5.950<br>[4.964, 6.937]<br>**p=0.000** | 62.063<br>[56.141, 67.984]<br>**p=0.000** | 739.866<br>[713.846, 765.885]<br>**p=0.000** |
| $\hat{\beta}_1$ | -0.021<br>[-0.050, 0.008]<br>p=0.153 | 0.065<br>[-0.058, 0.187]<br>p=0.298 | -1.288<br>[-2.104, -0.472]<br>**p=0.002** |
| $\hat{\beta}_2$ | -3.700<br>[-57.564, 50.164]<br>p=0.891 | -302.313<br>[-342.443, -262.183]<br>**p=0.000** | -1997.532<br>[-2666.820, -1328.245]<br>**p=0.000** |
| $\hat{\beta}_3$ | 0.057<br>[-0.763, 0.876]<br>p=0.891 | 4.185<br>[3.591, 4.780]<br>**p=0.000** | 26.300<br>[16.214, 36.386]<br>**p=0.000** |
| N | 70 | 70 | 70 |
| R<sup>2</sup> | 0.029 | 0.372 | 0.682 |

**Table A2.** Poisson Regression

| | $Y_1$ | $Y_2$ | $Y_3$ |
| --- | --- | --- | --- |
| $\hat{\beta}_0$ | 1.788 | 4.128 | 6.608 |
|  | [1.576, 2.001]<br>**p=0.000** | [4.065, 4.192]<br>**p=0.000** | [6.589, 6.626]<br>**p=0.000** |
| $\hat{\beta}_1$ | -0.004 | 0.001 | -0.002 |
|  | [-0.010, 0.002]<br>p=0.200 | [-0.001, 0.003]<br>p=0.256 | [-0.002, -0.001]<br>**p=0.000** |
| $\hat{\beta}_2$ | -0.770 | -7.152 | -4.730 |
|  | [-10.137, 8.596]<br>p=0.872 | [-10.321, -3.984]<br>**p=0.000** | [-5.740, -3.719]<br>**p=0.000** |
| $\hat{\beta}_3$ | 0.012 | 0.100 | 0.064 |
|  | [-0.129, 0.152]<br>p=0.871 | [0.053, 0.148]<br>**p=0.000** | [0.049, 0.079]<br>**p=0.000** |
| N | 70 | 70 | 70 |

95% CI in brackets. $p$ indicates the corresponding $p$ -value

**Table A3.** Quasi Poisson Regression

| | $Y_1$ | $Y_2$ | $Y_3$ |
| --- | --- | --- | --- |
| $\hat{\beta}_0$ | 1.788 | 4.128 | 6.608 |
|  | [1.559, 2.017]<br>**p=0.000** | [4.044, 4.213]<br>**p=0.000** | [6.554, 6.661]<br>**p=0.000** |
| $\hat{\beta}_1$ | -0.004 | 0.001 | -0.002 |
|  | [-0.010, 0.003]<br>p=0.238 | [-0.001, 0.003]<br>p=0.396 | [-0.003, -0.000]<br>**p=0.018** |
| $\hat{\beta}_2$ | -0.770 | -7.152 | -4.730 |
|  | [-10.854, 9.313]<br>p=0.881 | [-11.367, -2.938]<br>**p=0.001** | [-7.593, -1.867]<br>**p=0.002** |
| $\hat{\beta}_3$ | 0.012 | 0.100 | 0.064 |
|  | [-0.140, 0.163]<br>p=0.880 | [0.038, 0.163]<br>**p=0.003** | [0.021, 0.107]<br>**p=0.005** |
| N | 70 | 70 | 70 |

95% CI in brackets. $p$ indicates the corresponding $p$ -value

**Figure A1.**
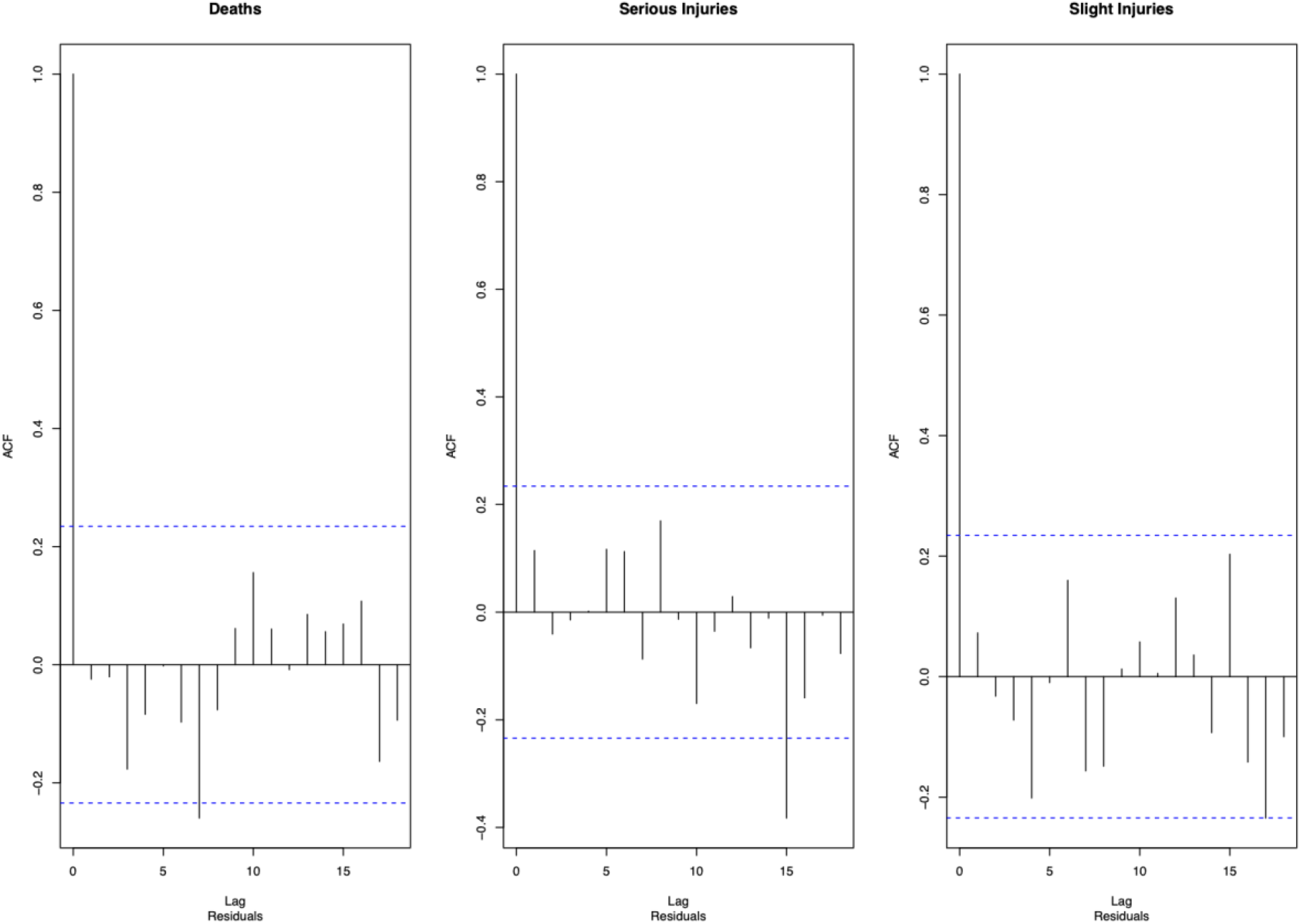
Regression residuals

**Figure A2.**
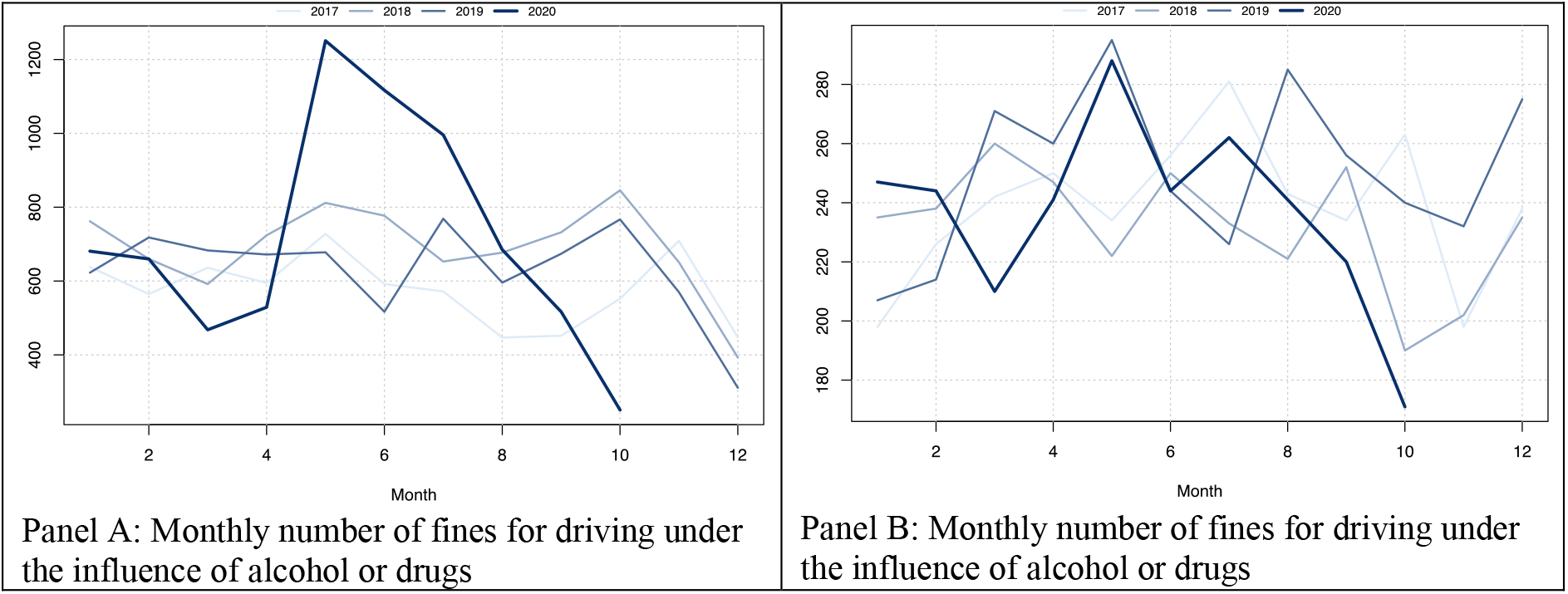
Monthly number of fines for speeding and driving under the influence of alcohol or drugs, years 2017-2020, Northern Ireland

